# C-reactive protein guided use of procalcitonin in COVID-19

**DOI:** 10.1101/2021.02.10.21251350

**Authors:** Rebecca Houghton, Nathan Moore, Rebecca Williams, Fatima El-Bakri, Jonathan Peters, Matilde Mori, Gabrielle Vernet, Jessica Lynch, Henry Lewis, Maryanna Tavener, Tom Durham, Jack Bowyer, Kordo Saeed, Gabriele Pollara

## Abstract

Low procalcitonin (PCT) concentrations (<0.5ng/mL) can facilitate exclusion of bacterial co-infection in viral infections, including COVID-19. However, costs associated with PCT measurement preclude universal adoption, indicating a need to identify settings where PCT provides clinical information beyond that offered by other inflammatory markers, such as C-reactive protein (CRP) and white cell count (WCC). In an unselected cohort of 299 COVID-19 patients, we tested the hypothesis that PCT<0.5ng/mL was associated with lower levels of CRP and WCC. We demonstrated that CRP values below the geometric mean of the entire patient population had a negative predictive value for PCT<0.5ng/mL of 97.6% and 100% at baseline and 48 hours into admission respectively, and that this relationship was not confounded by intensive care admission or microbiological findings. CRP-guided PCT testing algorithms can reduce costs and support antimicrobial stewardship strategies in COVID-19.

## Main text

Unnecessary antibiotic prescriptions during the COVID-19 pandemic will increase selection for antibiotic resistance (1). Optimising antimicrobial stewardship approaches in COVID-19 is a global priority, and a recent study has indicated that procalcitonin (PCT) <0.5ng/mL offered a high negative predictive value (>95%) for the presence of bacterial co-infections (2). Elevated PCT is associated more frequently with bacterial than viral infections (3), and thus low PCT can support cessation of antibiotic prescriptions in both COVID-19 and non-COVID-19 settings (4–8). Unlike the inflammatory markers C-reactive protein (CRP) and white cell count (WCC), PCT is not routinely measured (9), partly due to the costs associated with its testing (10,11). Elevations in PCT, CRP and WCC can be concordant (12), and therefore a key research question is to identify scenarios in which PCT provides clinical information beyond that offered by other inflammatory markers, in turn informing algorithms that avoid the expense of redundant PCT measurements.

We identified 299 adult patients with compatible clinical syndromes for COVID-19 and SARS-CoV-2 confirmed on molecular testing, admitted to Hampshire Hospitals NHS Trust between 5^th^ March and 26^th^ April 2020. The measurement of PCT, CRP and WCC at baseline and daily during hospital admission was standard of care. We tested the hypothesis that PCT concentration was closely related to CRP or WCC levels, choosing PCT≥0.5ng/mL as threshold for the presence of bacterial pulmonary infection (4,7,8,13,14). PCT≥0.5ng/mL was associated with greater levels of CRP on admission and 48 hours into admission, independent of the need for intensive care unit (ICU) admission during the hospital stay (figs 1A+C). In contrast, PCT≥0.5ng/mL was associated with only a modestly greater WCC in non-ICU attending patients (fig 1B+D). Next, we used pairwise comparison to test the hypothesis that low levels of CRP/WCC could exclude PCT≥0.5 ng/ml. We used the geometric mean of CRP or WCC for the entire patient population at each timepoint as cut-offs for low and high levels of these markers. Strikingly, low CRP was strongly associated with PCT < 0.5ng/mL (NPV 97.6% and 100% at baseline and 48 hours into admission respectively) (fig 1C). A similar relationship was observed for WCC, although with lower NPV (91.0% and 84.9% respectively) (fig 1D). Notably, patients with significant microbiological results, as previously defined (2), were not associated with elevated levels of any inflammatory marker (fig 1C+D) (9).

**Figure 1:**
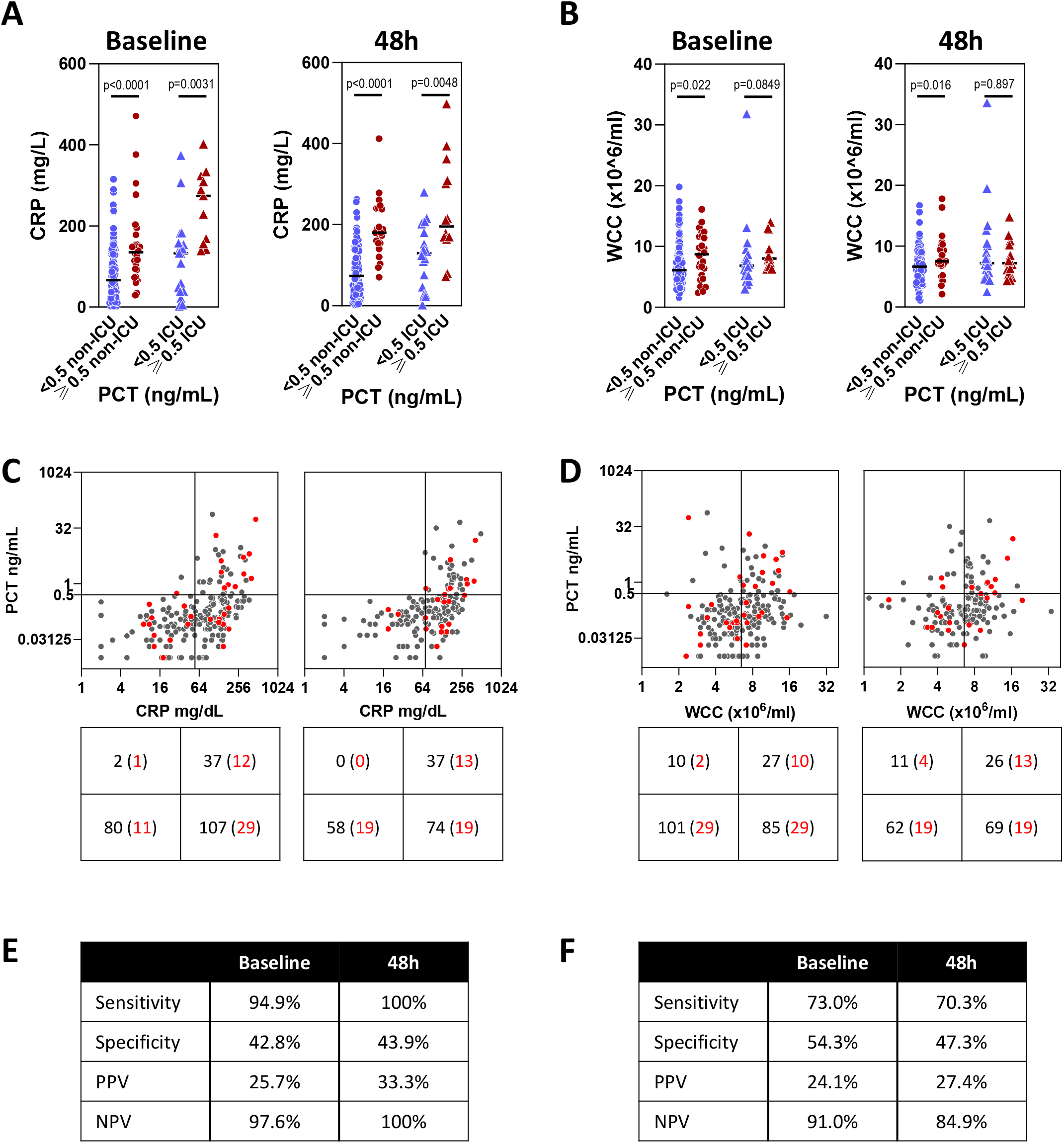
Association between CRP or WCC and the concentration of PCT in COVID-19. Concentration of CRP (A) or enumeration of WCC (B) stratified by PCT concentration <0.5 or ≥ 0.5ng/mL and by requirement for ICU admission during hospital stay. Assessments were made on admission to hospital (“baseline”) and 48 hours into hospital admission (“48h”). All p values derived using 2-tailed Mann-Whitney tests. Relationship between PCT and CRP concentration (C) or WCC (D). Scatter plot horizontal line represent PCT concentration cut-off (≥ 0.5ng/mL) and vertical line represents geometric mean of all patients at each timepoint, independent of PCT concentration. Total number of patients in each quadrant of scatter plot quantified in table below. Red dots and bracketed numbers reflect number of patients with significant microbiological findings. Sensitivity, specificity, PPV and NPV given for CRP (E) or WCC (F) to identify high or low PCT concentrations.

We demonstrate low CRP concentrations can predict low PCT values in COVID-19, precluding the cost of this test in approximately a third of patients. We acknowledge the retrospective, single-centre nature of the study, and the unspecified interval between symptom onset and hospital admission. However, these limitations are countered by the large, unselected populations, the consistent relationship between variables at both time points, and the independence from ICU admission status. The lack of association between PCT and microbiology results was notable and reminiscent of observations made in COVID-19 for other inflammatory markers (9). The diagnostic stewardship role for PCT in excluding bacterial co-infection at high CRP levels will need investigating through studies comparing COVID-19 with syndromically-defined bacterial pneumonia (9,15).

We declare no conflicts of interest.

## Data Availability

All anonymised data presented in the manuscript is available to others by request of the corresponding author.

